# Bead-assisted SARS-CoV-2 multi-antigen serological test allows effective identification of patients

**DOI:** 10.1101/2021.04.08.21254348

**Authors:** Yaiza Cáceres-Martell, Daniel Fernández-Soto, Carmen Campos-Silva, Eva M. García-Cuesta, Jose M Casasnovas, David Navas-Herrera, Alexandra Beneítez-Martínez, Pedro Martínez-Fleta, Arantzazu Alfranca, Francisco Sánchez-Madrid, Gabriela Escudero López, Carlos Vilches, Ricardo Jara-Acevedo, Hugh T. Reyburn, José M. Rodríguez Frade, Mar Valés-Gómez

## Abstract

Many new aspects of COVID-19 disease, including different clinical manifestations, have been identified during the pandemic. The wide array of symptoms and variation in disease severity after SARS-CoV-2 infection might be related to heterogeneity in the immune responses of different patients. Here we describe a new method for a simple multi-antigen serological test that generates a full picture of seroconversion in a single reaction. The assay is based on the detection by flow cytometry of multiple immunoglobulin classes (isotypes) specific for four SARS-CoV-2 antigens: the Spike glycoprotein (one of the highly immunogenic proteins), its RBD fragment (the major target for neutralising antibodies), the nucleocapsid protein and the main cysteine-like protease. Until now, most diagnostic serological tests measured antibodies to only one antigen and some patients seemed to not make any antibody response. Our data reveal that while most patients respond against all the viral antigens tested, others show a marked bias to make antibodies against either proteins exposed on the viral particle or those released after cellular infection. Combining all the four antigens and using machine learning techniques, it was possible to clearly discriminate between patients and healthy controls with 100% confidence. Further, combination of antigens and different immunoglobulin isotypes in this multi-antigen assay improved the classification of patients with mild and severe disease. Introduction of this method will facilitate massive screenings of patients to evaluate their immune response. It could also support vaccination campaigns both to select non-immune individuals and to distinguish infected patients from vaccine responders.

## INTRODUCTION

The novel single-stranded RNA-enveloped beta-coronavirus, called Severe Acute Respiratory Syndrome-Coronavirus-2 (SARS-CoV-2), causes the respiratory disease referred to as COVID-19 that was recognised by the WHO as a pandemic in 2020 [1, 2]. Average mortality ranges from 0.16 to 20.88% for women and 0.27–34.68% in men [3, 4], depending on age. Moreover, the virus spreads very quickly and a significant percentage of patients develop an exacerbated immune response with a widespread inflammation and multi-organ failure [5], which require long-term hospitalization and causes a serious problem to the health systems. Further, many cases are essentially asymptomatic, thus tests for viral infection are needed in order to follow the propagation worldwide and to identify the role of the virus, as new clinical manifestations are described in different patient groups.

As with any other infectious disease, antibodies are generated against certain viral proteins and detection of those antibodies can be used in diagnostics, to complement assays for viral nucleic acids and to follow the evolution of the infection. One of the major antigens of SARS-CoV-2 is the envelope Spike (S), which mediates attachment to host cells and virus cell entry via its Receptor Binding Domain (RBD) [6, 7]. Antibodies directed against the RBD can often neutralise the infection [8]. Other viral antigens include the nucleocapsid protein (NP) and the 3CL main protease (Mpro), which are only synthesised once the virus has infected the cell. These viral proteins also generate antibody responses [7, 9] and can be used in serological tests [9, 10]. In fact, antibodies to the viral protease could be detected in plasma and saliva of individuals who had been infected with SARS-CoV-2 [10].

Antibody tests to detect exposure to SARS-CoV-2 are commercially available in several formats, such as ELISA, CLIA and lateral flow devices. These assays are useful tools for epidemiological studies that need to identify infected people. However, the commercial assays usually test for antibodies to only one antigen, generally either Spike or the nucleocapsid protein. As the pandemic advances and new clinical manifestations are described, it is important to evaluate the quality (e.g. antibody isotypes, response to different antigens), quantity (antibody titre) and duration of the immune response in patients with different severity and symptoms. Carrying out multiple ELISA assays, to analyse several antigens and immunoglobulin types over plasma dilutions for each individual, greatly increases the amount of reagents and time needed to evaluate large cohorts of patients. This thorough immune characterization would be of particular interest to follow up vaccination efficacy, as well as to follow up certain population groups such as immunodeficient patients or other high risk individuals.

To facilitate the implementation of screenings of COVID-19 patient populations, we here report the development of a multiplex bead-based flow cytometry assay that assesses, in a single reaction, for sero-reactivity to four different SARS-CoV-2 antigens: the S protein extracellular region, its RBD, NP and Mpro. This test includes the analysis of IgA, IgG, IgM, which can also be performed simultaneously by using different fluorophores for each anti-isotype on any standard flow cytometer (488nm or 633nm excitation), and do not require the use of specialized software. The technique yields results with extremely low background signals and has specificity and sensitivity near 100%, therefore providing a very good tool to have a full view of COVID-19 patient immune response. Differences in the specific Ig responses against the four antigens allows easy discrimination between vaccinated and naturally-infected individuals. Further, machine learning analysis allowed classification of patients and healthy controls, without any error just using IgG data. Thus, a simple multi-antigen serological test clearly discriminates between patients and healthy controls with a 100% confidence. Modification of the algorithm to take Ig isotype data into account also allowed high confidence discrimination between mild and severe presentations of COVID-19 disease, at least retrospectively.

## METHODS

### Patient selection, samples and Institutional Review Board permits

Experiments were carried out following the ethical principles established in the Declaration of Helsinki. Patients (or their representatives) were informed about the study and gave a written informed consent. This study used samples from several hospitals. For optimization experiments, samples from the research project “Immune response dynamics as predictor of COVID-19 disease evolution. Implications for therapeutic decision-making” approved by La Princesa Health Research Institute Research Ethics Committee (register # 4070) were used; samples and data from patients with severe vs mild disease were provided by the Biobank Hospital Universitario Puerta de Hierro Majadahonda (HUPHM)/Instituto de Investigación Sanitaria Puerta de Hierro-Segovia de Arana (IDIPHISA) (PT17/0015/0020 in the Spanish National Biobanks Network), they were processed following standard operating procedures with the appropriate approval of the Ethics and Scientific Committees. For comparison of disease severity, 29 COVID-19 patients, diagnosed by PCR, were recruited. 14 patients, classified as mild disease or asymptomatic, did not require treatment after diagnosis. 15 patients, classified as severe disease, required ICU hospitalization (Supplementary Table 1). Plasma samples were obtained 33-40 days after diagnostic PCR and, separated by blood centrifugation after collection in EDTA tubes, 15 plasma samples collected from healthy blood donors before June 2019 (PRE-COVID-19) in the Puerta de Hierro hospital biobank, were used as negative controls.

15 vaccinated (Pfizer BioNTech) individuals were recruited at the Centro de Hemoterapia y Hemodonación de Castilla y León (ChemCyL) for comparison with SARS-CoV-2 infected patients (Supplementary Table 2). These samples were obtained as part of the project “Development of serological assays for detection of viral antigens (SARS-COV2)”. The protocol was approved by the Bioethics Committees: CEIm Área de Salud Valladolid Este, Hospital Clínico Universitario de Valladolid, with the number/BIO 2020-98-COVID.

CPD respiratory panel human plasma samples were obtained from a commercial source (BioIVT - West Sussex, United Kingdom).

### Expression of the SARS-CoV-2 Cys-like protease (Mpro), nucleocapsid (NP), Spike (S) and RBD proteins

Recombinant SARS-CoV-2 proteins were expressed with a histidine tag. Cys-like protease (Mpro) and nucleocapsid (NP) proteins constructs were expressed in the *E. coli* strain BL21 Star (DE3) pLysS (ThermoFisher) and purified as described [10].

Recombinant cDNAs coding for soluble S (residues 1 to 1208) and RBD (332 to 534) proteins were cloned in the pcDNA3.1 vector for expression in HEK-293F cells using standard transfection methods. The two constructs contained the S signal sequence at the N-terminus, and a T4 fibritin trimerization sequence, a Flag epitope and an 8xHis-tag at the C-terminus. In the S protein, the furin-recognition motif (RRAR) was replaced by the GSAS sequence and it contained the A942P, K986P and V987P substitutions in the S2 portion. Proteins were purified by Ni-NTA affinity chromatography from transfected cell supernatants and they were transferred to 25 mM Hepes-buffer and 150 mM NaCl, pH 7.5, during concentration.

### Bead based flow cytometry assay for detection of antibodies to SARS-CoV-2

10^6^ magnetic fluorescent beads, with a mean diameter 5.5 µm and high density carboxyl functional groups on the surface (QuantumPlex™M COOH – Bangs Laboratories, Inc.), were covalently coupled with 30 µg of viral protein through their primary amines by two-step EDC/NHS protocol. Beads were resuspended in a solution of PBS containing 1% casein and a stabilizer (Biorad 1x PBS blocker). To distinguish the beads coated with different antigens, different fluorescence intensity combinations in the APC and PerCP channels were used (Figure 1A).

**Figure 1.**
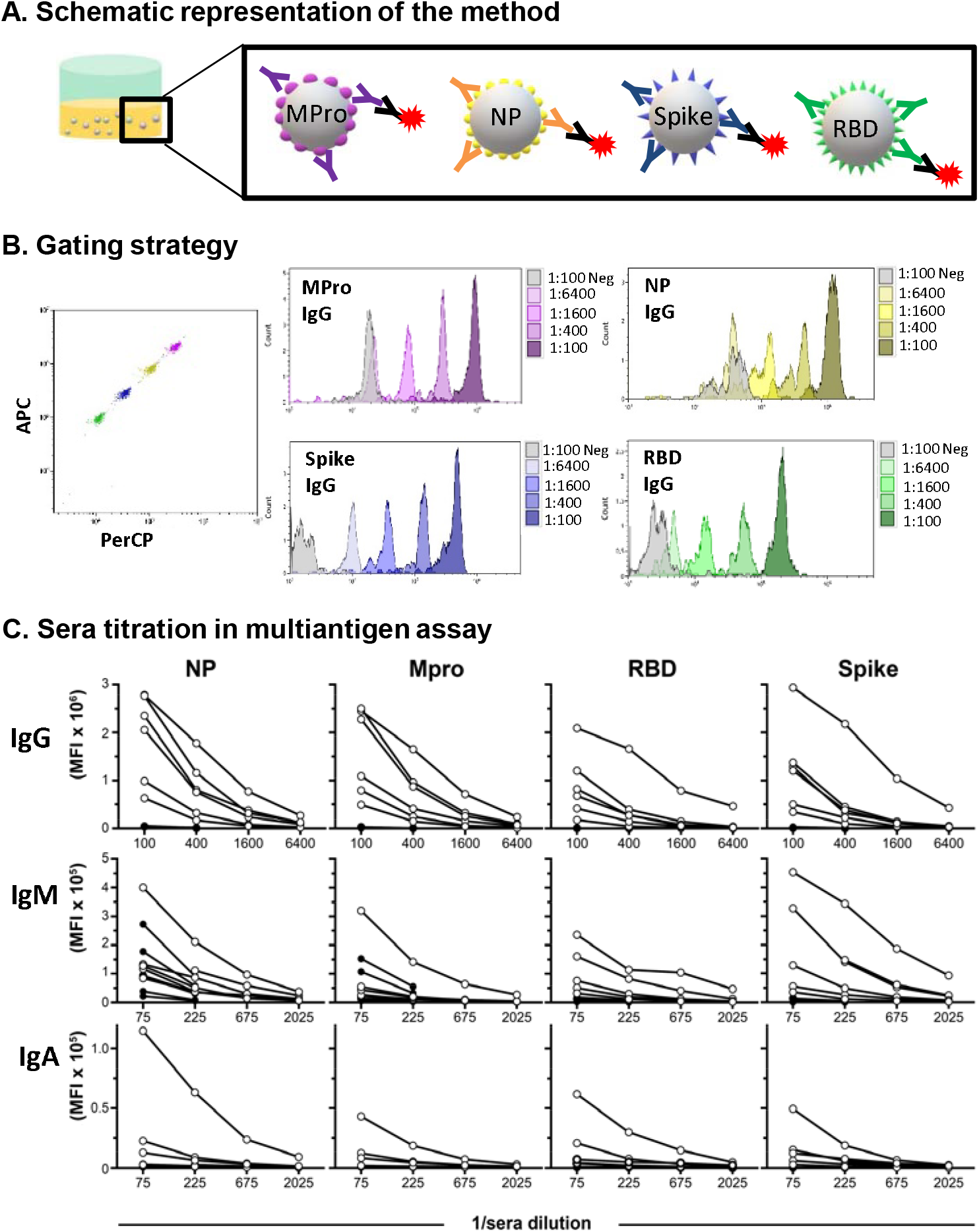
A basic bead-assisted multiantigen assay for antibody detection in COVID-19 human serum samples. **A. Schematic representation of the method.** Four different SARS-CoV-2 His-tagged antigens (Mpro, NP, S and RBD) were covalently coupled to magnetic beads labeled with dyes showing different fluorescence intensity in the APC and PerCP channels. Equal amounts of the different bead populations were mixed in the same tube and incubated with dilutions of plasma from patients or healthy donors, as indicated. Antibodies bound to the antigen were developed with fluorophore-conjugated anti-human Ig and samples were analysed by flow cytometry. **B. Gating and antibody detection strategy**. Magnetic beads coupled with individual SARS-CoV-2 antigens were mixed in a single well and incubated with the indicated dilutions of plasma from a healthy donor and a COVID-19 patients. Subsequently, detection was performed in two separate tubes, one with PE-conjugated anti-human IgG and the second tube containing PE-conjugated anti-human IgM + FITC-conjugated anti-human IgA. The FSC/SSC region corresponding to 6 µm beads was selected and individual populations of beads were visualized in a APC/PerCP dot plot (left). Antibody bound to each bead type was analyzed independently in histograms within each bead gate. The plots represent Mean Fluorescence Intensity (MFI) values from the analysis of IgG obtained for the individual bead regions of one patient, comparing with a negative control (pre-COVID-19). **C. Sera titration in the multi-antigenic assay**. Antibody MFI values obtained for 6 healthy donors (closed symbols) and 6 COVID-19 patients (open symbols) in a multiantigen assay including a range of serum dilutions, as described in B.

Beads were incubated with either rabbit anti-His-tag antibody (Proteintech Group) or plasma from patients or healthy donors in a final volume of 50 μl in 96-well-plates (Nunc™ MicroWell™ 96-Well, Thermo Fisher Scientific) using the dilutions indicated in each experiment. Patient plasma samples were diluted in PBS-casein (Biorad,1x PBS blocker), and incubated with the beads for 40 min at room temperature under agitation. Beads were washed three times by addition of PBS, placing the tubes or plates on a magnet (MagneSphere® Mag. Sep. Stand 12-hole, 12×75mm, Promega; Handheld Magnetic Separator Block for 96 well plate, Merck, Millipore) and decantation of supernatant.

To visualize antibody bound to antigen-coated beads, either PE-conjugated anti-rabbit antibody (0.25 μg/ml, Southern Biotech), PE-conjugated anti-human IgG and IgM, or FITC-conjugated anti-human IgA antibody (Immunostep S.L.) were added (30 μL/well) and incubated for 20 minutes at room temperature under agitation. After three washes, data were acquired by flow cytometry using either CytoFLEX or Cytomics FC 500 (Beckman Coulter).

For large screenings performed in different days, data were normalized to the values of a positive control serum included in every assay.

### ELISA for detection of antibodies to SARS-CoV-2

ELISA assays for detection of antibodies directed against the four SARS-CoV-2 antigens were carried out as described [10].

### Statistical analysis

To assess the prediction capacity of the new methodology, an algorithm was built using Scikit-learn python package [11] (code available on request). Samples were stratified and randomly spliced into a training and a test set. The training samples were used to fit a random forest classifier which then predicted the healthy vs disease category of unseen test samples (1/7 of total samples). This was repeated n=10,000 times. For each patient, accuracy was calculated as the proportion of correct predictions divided by the number of predictions made. As a complementary approach, a mean Receiver Operating Characteristic (ROC) curve was built for the random forest classifier by stratified 15-fold cross-validation, using the smaller set (2-3 samples) to train the model and then predicting the remaining ones.

For heatmap representation, each variable was scaled to a range (0,1) using the *MixMaxScale*r command from *Scikit-learn* and visualized using *heatmap* command from *seaborn* python packages. For Principal Component Analysis, each variable was scaled as described, and the *PCA* command from *Scikit-learn* was used to fit and transform the data. Principal components up to a 95% of accumulated explained variance were saved.

Comparison between severe and mild patients in each variable was performed by multiple t-tests followed by False Discovery Rate (1%) correction by two-stage step-up method in Graph Pad Prism 8 Software (GraphPad Software, USA, www.graphpad.com).

## RESULTS

### Basic bead-assisted multi-antigen serological assay using flow cytometry

The NP, S and RBD proteins of Coronaviruses have been widely used in single-antigen serological assays for SARS and MERS-caused diseases [12]. However, the use of these antigens in combination with the immunogenic Mpro [10], can more fully describe the magnitude and duration of the immune response in SARS-CoV-2-infected patients. In order to facilitate comprehensive characterization of COVID-19 patients with a high throughput approach, a multi-antigen assay was developed with several viral antigens immobilised on fluorescent beads, to allow flow cytometry detection of the multiple antibodies generated during SARS-CoV-2 infections.

As depicted in Figure 1A, the assay uses fluorescent magnetic beads coated with SARS-CoV-2 antigens. Each protein was immobilised on a bead population with a particular fluorescence intensity in the red channel (e.g. APC/PerCP). This permits simultaneous detection of antibodies to different antigens in a single test tube or well. After incubation with patient plasma, one or several secondary antibodies conjugated to different fluorophores, such as FITC or PE, were used for identification of the IgG, IgA and IgM immunoglobulins bound to the viral antigens. In most experiments, combinations of anti IgM-PE and IgA-FITC were used. IgG and IgA were also combined with good results. Thus, the data for each antigen-specific Ig could be determined in a single reaction, by using three different fluorophores. Specifically, in several experiments the FITC, PE and PE-Cyanine7 fluorochrome combination was tested for the detection of IgG, IgA and IgM respectively.

Initially, to define the detection limits and the amount of antibody binding, titration experiments varying the amount of beads (not shown) and the concentration of anti-His-tag antibody were performed (Supplementary Figure 1). These trial experiments allowed estimation of the signal for a known concentration of antibody and the data suggested that the new methodology could provide good sensitivity. Indeed, since all the antigen constructs have only a single His-tag, it would be expected that the use of plasma containing a polyclonal mixture of antibodies binding multiple epitopes would provide more signal and further increase sensitivity.

The anti-His signal obtained for the S protein was lower compared to other viral antigens, but did not affect detection in patient plasma. The lower detection of S by His-tag antibody was likely due to a lower molar amount of S than NP, MPro or RBD bound to the beads. This was expected because S molecular weight (∼180 KDa) is at least four times higher than the other antigens (25-40 KDa). Sera analysis allowed a very good separation of control and convalescent samples in a wide range of dilutions (Figure 1B,C). The multi-antigen assay also yielded good results when combining three anti-human secondary antibodies conjugated to different fluorophores (Supplementary Figure 2). Thus, the use of magnetic beads and flow cytometry is a suitable technique for the serological analyses.

**Figure 2.**
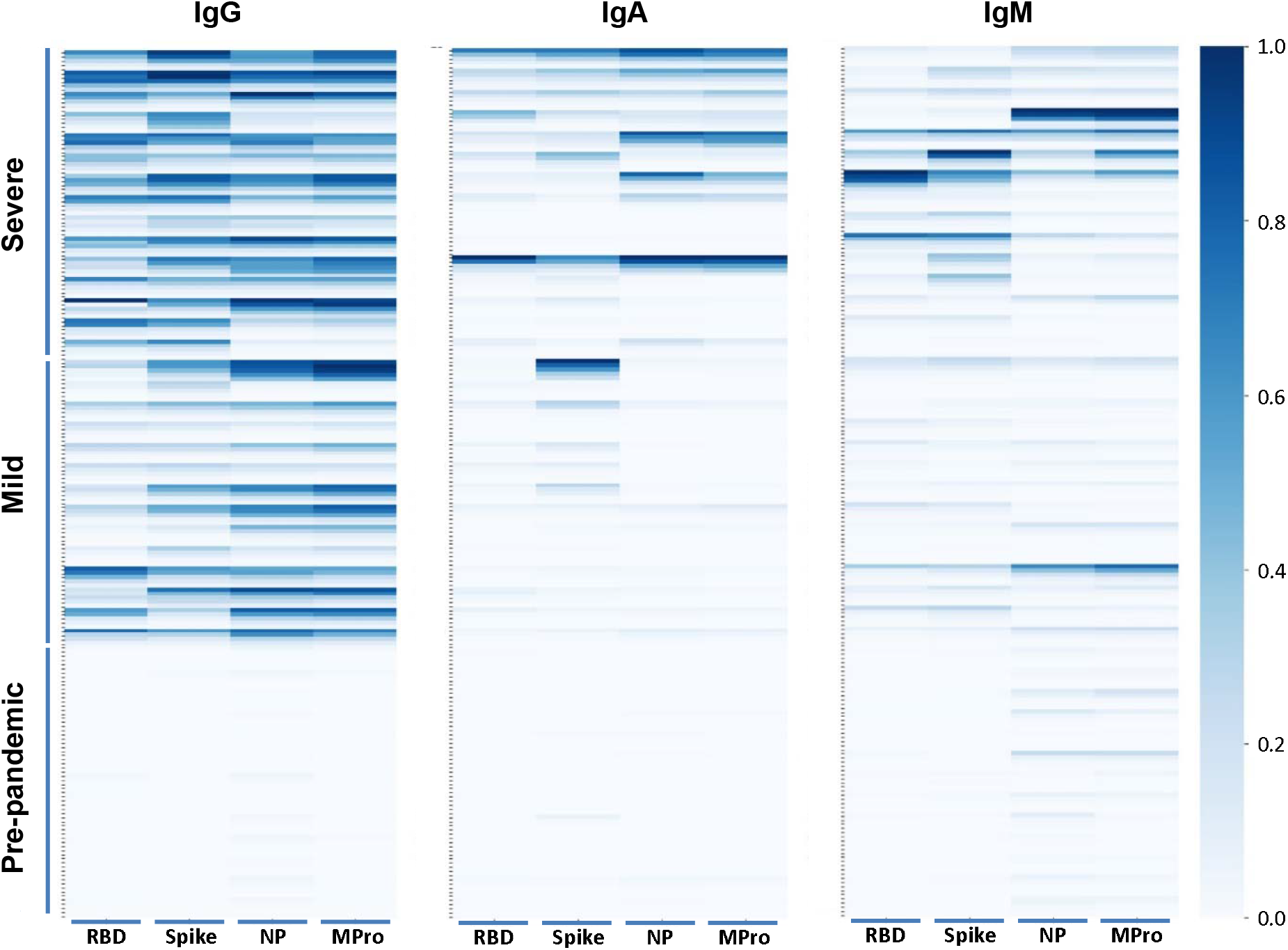
Heat map representing antibody titers from multi-antigen COVID-19 assays. Sera from 15 healthy controls and 29 COVID-19 patients were incubated with four different SARS-CoV-2 antigens coated beads: S, RBD, NP, and Mpro (indicated at the bottom), detected with antibodies to identify IgG, IgA and IgM and analysed by flow cytometry using the multi-antigen assay described in Figure 1. To summarize all the data, a heat map was built showing the intensity of the IgG/IgM/IgA antibodies detected in donor sera. Each column corresponds to one antigen and rows include five different dilutions (1/100,1/200, 1/600, 1/1800, 1/5400) for each individual. The intensity of the blue color depicts the amount of antibody.

### Multi-antigen bead-assisted flow cytometry identifies COVID-19 patients with 100% confidence

The sensitivity and specificity of the new method was evaluated by testing for the presence of antibodies against four SARS-CoV-2 antigens (S, RBD, NP, Mpro) in 44 plasma samples, including 29 COVID-19 patients, 14 of them with mild disease and 15 with severe disease. Each plasma sample was tested over a range of dilutions (1:100 to 1:5400) for three Ig isotypes (IgA, IgG, IgM). Initial analysis using heat map representations of the data (Figure 2), shows a clear difference between the signal obtained for IgG antibodies against the four antigens between healthy controls and COVID-19 patients. As expected, although, IgA and IgM SARS-CoV-2-specific antibodies were detected in multiple patients, they were not present in all the sera tested. While IgM had a higher background, IgA provided very clean and specific data. To investigate the analytical specificity, the possible cross-reactivity of antibodies against other microorganisms that produce symptoms of respiratory disease was analysed. Samples from 16 patients characterised as IgG positive for the following microorganisms were selected: MERS-CoV, H. Influenzae, RSV, Influenza A, Influenza B, Parainfluenza, Adenovirus, Enterovirus, M. pneumoniae, Legionella, C. pneumonia. These data show that other respiratory infections do not generate SARS-CoV-2 reactive antibodies (Supplementary Figure 3).

**Figure 3.**
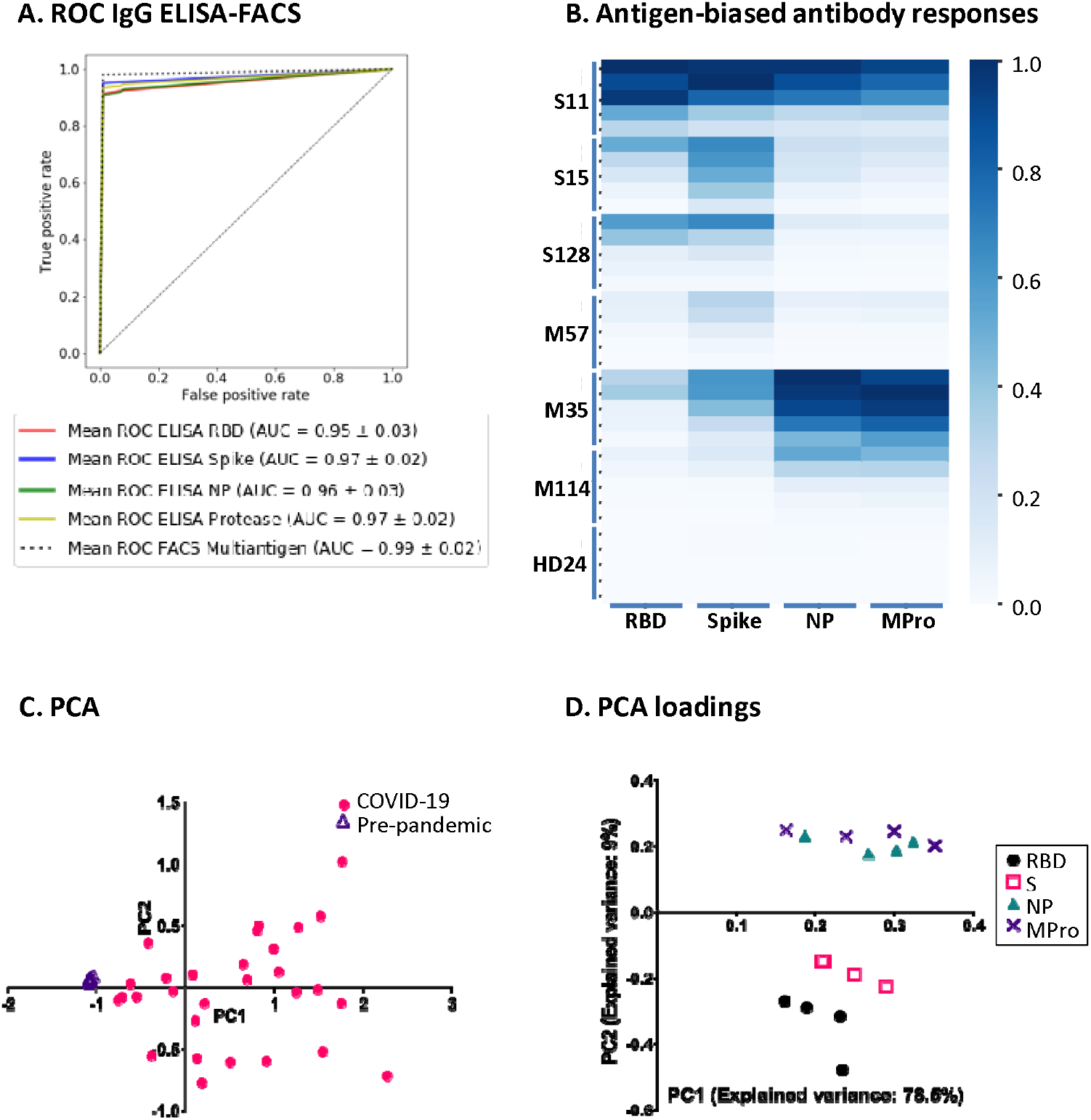
Multi-antigen serological assay identifies COVID-19 patients with nearly 100% confidence. **A. Receiver Operating Characteristic (ROC) curves of single-antigen ELISA and multi-antigen FACS assays.** A random forest classifier was trained with one healthy and 2 COVID controls IgG values and used to predict the rest of the samples. The mean ROC curve after 15-fold cross-validation is shown for each condition. **B. Heat map of patients with biased IgG response against one type of viral antigens**. Although most COVID-19 patients respond by producing antibodies against the four antigens tested, 5 out of 29 donors responded preferentially to either S/RBD or NP/Mpro. Data from 6 patients and 1 healthy donor are shown for comparison. **C. Principal components Analysis (PCA)**. A principal component analysis was run with data from IgG antibodies (dilutions 1/100,1/200, 1/600, 1/1800), and the two first principal components were used to represent each patient. Triangles and circles represent pre-pandemic controls and COVID-19 patients, respectively. **D. PCA loadings**. Visual representation of the loadings of the two first principal components of the PCA. Each dilution of IgG titer against RBD, Spike, NP and MPro (as indicated) is represented as a separate variable.

Machine learning techniques were used to assess the specificity and sensitivity of this novel methodology. A random forest classifying algorithm was developed to evaluate the prediction capacity of seronegative versus COVID seropositive individuals when data were generated by ELISA and by FACS, comparing both single-antigen and multi-antigen techniques. When only the data generated for IgG by FACS were analysed, all the patients were correctly classified in 100% of the multi-antigen repetitions, except for two that were correctly classified 99.93% and 98.24% of the times (Supplementary Table 3). Combining data for the four antigens and 4 dilutions for IgG provides an overall prediction capacity of 99.94% true positive rate and 100% true negative rate. True positive rates near to 100% were also obtained when only three antigens (RBD, S and Mpro) and one dilution were analysed, highlighting the predictive power of the technique [IgG 1/100 99.87; IgG 1/200 99.98; IgG 1/600 98.94; IgG 1/1800 x=99.98]. In all cases, true negative rate was always 100%. Individual antigens by ELISA had slightly lower prediction values. Thus, the use of a single test including three antigens, one isotype detection (IgG) and one dilution results in accurate classification of patients, facilitating large screenings.

### COVID-19 patients respond differentially to the four viral antigens

Using a small training set, ROC curves were generated to compare the sensitivity and specificity of each single-antigen ELISA test and for the multi-antigen FACS technique (Figure 3A), and the latter again demonstrated the best performance, highlighting that a multi-antigen approach could be more useful in clinical contexts in which a high number of unknown samples must be classified using a limited amount of known controls.

The enhanced efficiency of the multi-antigen test is likely related to the observation that some patients clearly respond preferentially to antigens present in the viral particle (S, RBD), while other patients respond mainly to antigens normally only exposed once cells have been infected (NP, Mpro) (Figure 3B) [10]. The existence of this bias was independently confirmed when a Principal Component Analysis (PCA) was performed with data for each antibody isotype. This analysis revealed a clear separation of seropositive and seronegative patients (Figure 3C). Inspection of the PCA loadings (Figure 3D) showed that, for IgG, the second principal component discriminated between production of antibodies against either NP+MPro or S+RBD. Similar patterns were noted when IgA and IgM responses were analysed (not shown). The detection of this bias when analysing only a limited number of patients suggests that preferential antigen-specific responses are common and makes a strong case for the use of multi-antigen serological assays to avoid false-negative results. As the pandemic has advanced, it has been established that not all the patients respond in the same manner to the infection by SARS-CoV-2. In fact, a large body of clinical manifestations have been described and it has been suggested that different types of immune response may contribute to these different presentations. Therefore, it will likely be relevant to characterise potentially biased antibody responses when exploring the association between SARS CoV 2 infection and different clinical manifestations.

In aggregate, the multi-antigen assay produced data that easily and efficiently discriminated between seronegative and COVID seropositive individuals.

### Multi-antigen, multi-isotype analysis of COVID-19 patients antibody response improves classification related to disease severity and allows discrimination between vaccine-induced and naturally infected antibody responses

In general, patients with higher antibody titres are more likely to have suffered a severe infection, indicating infection severity is linked to increased antibody titres [13]. However, analysis of only IgG responses did not clearly discriminate between patients who had suffered severe or mild disease (Figure 2). Statistically significant increased antibody responses in severe compared to mildly-affected patients were observed in the case of IgA antibodies against NP (dilutions 1:100-1:600), MPro (dilutions 1:100-1:600) and RBD (dilutions 1:100-1:200). Using these variables to build a random forest allowed a classification into mild vs severe disease with a 92% accuracy (Figure 4) when IgG data (dilutions 1:600-1800) and the IgA (dilution 1:100) responses were analysed simultaneously, compared to an accuracy of 90% when only IgG data is taken into account (Figure 4). Although the analysis of a greater number of data is required, our results suggest the importance of analysing the IgG/IgA immune response against multiple SARS-CoV-2 antigens to establish clear criteria for severity discrimination.

**Figure 4.**
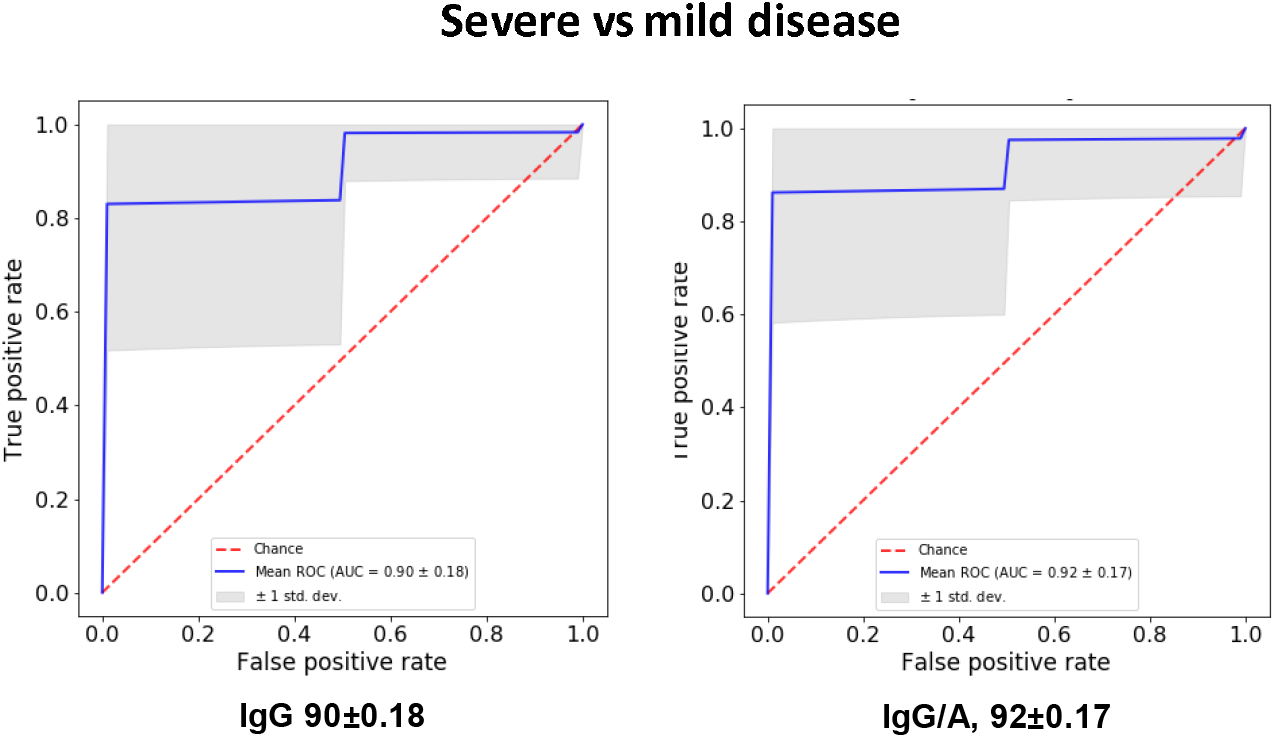
ROC curves classifying COVID-19 patients as either severe or mild disease. A random forest was trained to discriminate between COVID-19 patients with either severe or mild disease, using either IgG data alone or including data from other isotypes, and then used to predict unseen patients (1/7 of total samples). The mean ROC curve after 300 random repetitions is shown for each condition.

Comparison of the IgG and IgA responses against the four SARS-CoV-2 antigens were studied in samples from 15 vaccinated individuals and compared with that of naturally-infected individuals (Supplementary Table 2). As expected, vaccinated individuals only showed antibodies reactive with the S and RBD antigens, and IgG was the predominant isotype in vaccinated donors (Figure 5), while sera from naturally infected donors presented antibodies against all four viral antigens. Interestingly, only minimal IgA responses were observed in vaccinated donors, in contrast with naturally infected individuals who showed high IgA titres.

**Figure 5.**
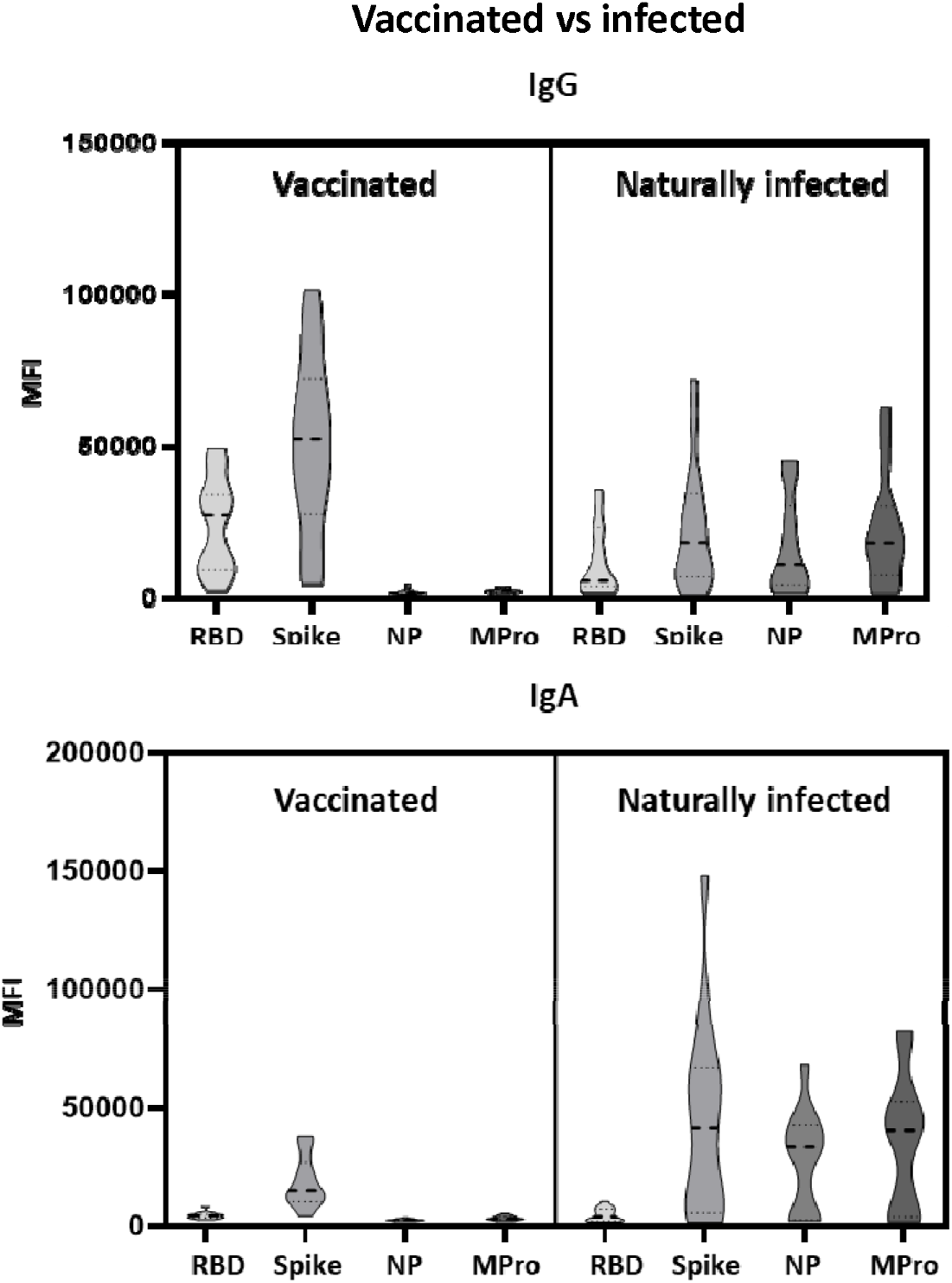
Multi-antigen serological shows a milder IgA response in vaccinated individuals compared to COVID-19 convalescents. Plasma IgG and IgA from 15 vaccinated donors (Pfizer BioNTech) were analized by flow cytometry using the multi-antigen assay described in Figure 1. Plots represent the comparison of the immunoglobulins produced against each individual antigen by each group of donors, vaccinated and SARS-CoV-2 naturally infected individuals.

## DISCUSSION

The development of fast, sensitive serological assays to detect exposure to SARS-CoV-2 is important. Unlike tests based on the detection of viral nucleic acids, serological tests detect antibodies which remain in serum after elimination of the virus and recovery from disease [14]. Current serological tests include classical ELISA techniques, serum chemiluminescence immunoassays (CLIA) against viral proteins and IgG/IgM lateral immunochromatography. Commercial presentations of these tests usually involve the detection of antibodies to only one viral antigen, generally S or NP [9, 15].

Here we report the development of a robust, quantitative, multiplex methodology that provides a much more complete description of the humoral immune response to infection with SARS-CoV-2 with excellent sensitivity and specificity. The method can be easily put into practice in most hospitals and clinical laboratories. The simultaneous detection of antibodies to multiple viral proteins in a single tube greatly facilitates sample handling and comparison of the antibody response to different viral antigens, a consideration that is even more important given the identification of COVID-19-convalescent patients whose antibody responses appear to be strongly biased for specific viral antigens. Indeed, patients that only respond to RBD or Spike would be classified as dubious or seronegative if a test for NP antibodies only was used. Similarly, a test for Spike antibodies would not identify patients that respond only to antigens released from infected cells, such as the NP and Mpro. IgM was only present in certain patients, as it corresponds to earlier disease stages. Thus, the multi-antigen test essentially eliminates the problems of false negatives and positives, even in low-seroprevalence settings [16].

In this study, we confirmed our previous data, describing a strong correlation between the antibody responses against intracellular antigens like NP and Mpro [10]. Further, while the majority of patients produced antibodies against all the antigens tested, we also identified several individuals who made responses with a marked bias for either antigens exposed on the viral particle envelope (S, RBD) or those that are mainly intracellular (P, NP). These data suggest distinct humoral immune responses among individuals, perhaps depending on the cellular damage caused by the virus infection. Importantly, serological assays based on the detection of antibodies to only one viral antigen are unlikely to detect people with strongly biased immune responses. The use of multiplex assays will help to understand the significance of this biased response.

Since the test can be automated and performed in a single reaction, the technique described here permits a simple, rapid and complete serological analysis of many patients, as it can be performed in multi-well plate compatible flow cytometers. In addition, the amount of protein required to coat beads is lower than that required for ELISA. All the advantages from this novel method could be of considerable importance to support vaccination campaigns and to select convalescent plasma for therapeutic use. Further, the use of antigens that are not part of the vaccine formulation will allow discrimination between those individuals responding to the vaccine from those with antibodies to other viral antigens, because of infection either shortly before or after vaccination (Figure 5). Finally, accurate serological testing will be critical to monitor the duration of the immune response after vaccination and to discriminate between vaccine-mediated protection from infection or disease.

Moreover, this assay has been designed to be easily put into practice in clinical settings and it would be straightforward to include more viral antigens as they are discovered to be immunogenic, and so further optimise the sensitivity and predictive value of the serological analyses in different clinical settings. The simultaneous analysis of immunoglobulin isotypes and multiple antigens in one reaction can be combined with automated data analysis, allowing rapid evaluation of serological status.

In this work we also tested the differences in serological responses between patients with mild and severe disease. Using the algorithms described here, patients requiring ICU hospitalization could be discriminated, with high confidence, from patients who experienced mild disease, something that appears difficult to achieve when the antibody response to only one antigen is assayed. It will be interesting to analyse the usefulness of these predictions on a prospective basis in order to evaluate its ability to suggest prognosis.

The assay reported here also has great potential to facilitate thorough analyses of serological responses from patients with different SARS-CoV-2 disease manifestations. The multi-antigen test can provide results in large screenings to aid in testing for a correlation between a given antibody response and a clinical aspect. In addition to an impact on early classification of patients, current limitations in the availability of vaccine doses suggest a novel possible application for sensitive multi-antigen assays for SARS-CoV-2 seropositivity. It has been shown that the antibody response to the first vaccine dose in individuals with pre-existing immunity is comparable or greater to that observed in naïve individuals who have been immunized twice [17]. Screening of the unvaccinated population with an assay sufficiently sensitive to identify individuals previously infected despite waning of antibody titres over time, would allow these individuals to be given the vaccine as a single booster, sparing them from possible suffering and complications after a second dose, and freeing up many urgently needed vaccine doses to be given to individuals with no protection. The test described here would also provide comprehensive information to support selection of convalescent sera or plasma for therapeutic use. Our data indicate the importance of this multi-antigen, multi-isotype analysis to detect potential SARS-CoV-2 reinfections in vaccinated individuals and suggest a possible use in establishing alternative vaccine administration routes that may elicit more potent IgA responses.

This multi-antigen and multi-Ig assay can be easily modified for detection of antibodies in other fluids as saliva and breast milk. It is also highly tunable to different research needs, including detection of different immunoglobulins, other viral proteins and even other potential antigens present in vaccine formulations.

In conclusion, the highly sensitive multi-antigen assay for flow cytometry offers the possibility of performing screenings on antibody response against SARS-CoV-2 both for research and to support patient diagnosis and development of therapeutic approaches based in convalescent plasma. Importantly, it provides a tool for the follow up of vaccinated individuals.

## Data Availability

All data are available upon request

## Acknowledgements

The authors would like to thank the director of the CNB-CSIC, M. Mellado, for coordination; the donors and the Biobank Hospital Universitario Puerta de Hierro Majadahonda (HUPHM)/Instituto de Investigación Sanitaria Puerta de Hierro-Segovia de Arana (IDIPHISA) (PT17/0015/0020 in the Spanish National Biobanks Network) for the human specimens used in this study; the Infectious Diseases and Intensive Care Units of HUPHM, for organization of sample and clinical data collection; Sofía Garrido and Miriam García, from the Immunology Dept. of HUPHM, for volunteering to process blood samples in difficult circumstances; the Centro de Hemoterapia y Hemodonación de Castilla y León (ChemCyL) for sample collection. This work was supported by the Spanish National Research Council (CSIC, project number 202020E079 and CSIC-COVID19-028) and grants from Madrid Regional Government “IMMUNOTHERCAN” [S2017/BMD-3733-2 (MVG)]; the Spanish Ministry of Science and Innovation [(MCIU/AEI/FEDER, EU): RTI2018-093569-B-I00 (MVG), SAF2017-82940-R (JMRF), SAF2017-83265-R (HTR); SAF2017-82886-R (FSM)]; Health Institute Carlos III (ISCIII) [RETICS Program RD16/0012/0006; RIER (JMRF); PI19/00549 (AA)]. The study was also funded by “La Caixa Banking Foundation” (HR17-00016 to FSM) and Fondo Supera COVID (CRUE-Banco de Santander) to FSM.

## Author contributions

YCM, DFS, CCS, EMGC, DN, AB, PMF, AA, JMRF, MVG, JMC, HTR prepared reagents, implemented experiments and analysed data; designed and optimized ELISA experiments; PMF, AA, FSM, GEL, CV selected patients and performed clinical evaluation; DFS carried out statistical analysis; RJA, JMRF, MVG, FSM, HTR were responsible for the conception and design of the study and obtaining financial support; DFS, HTR, JMRF, MVG, wrote the manuscript with revisions from all authors.

