## Supplemental Figures and tables for "Bead-assisted SARS-CoV-2 multi-antigen serological test allows effective identification of patients"

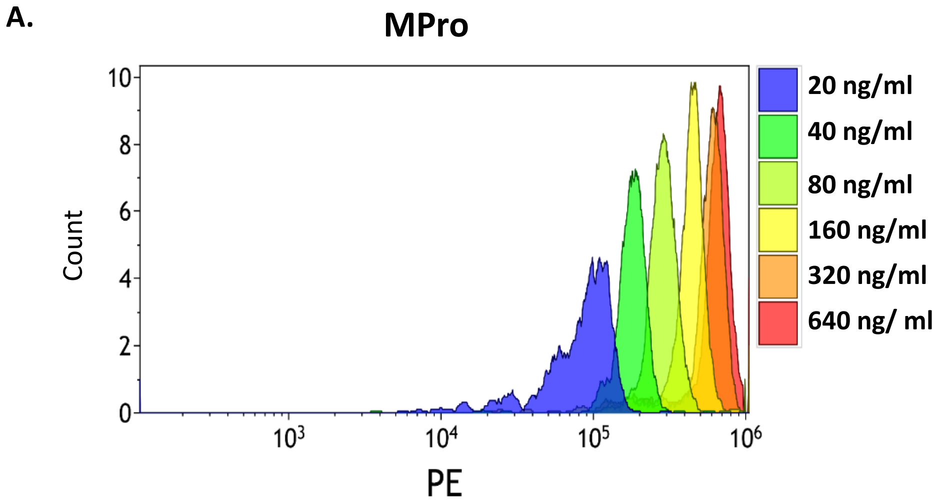


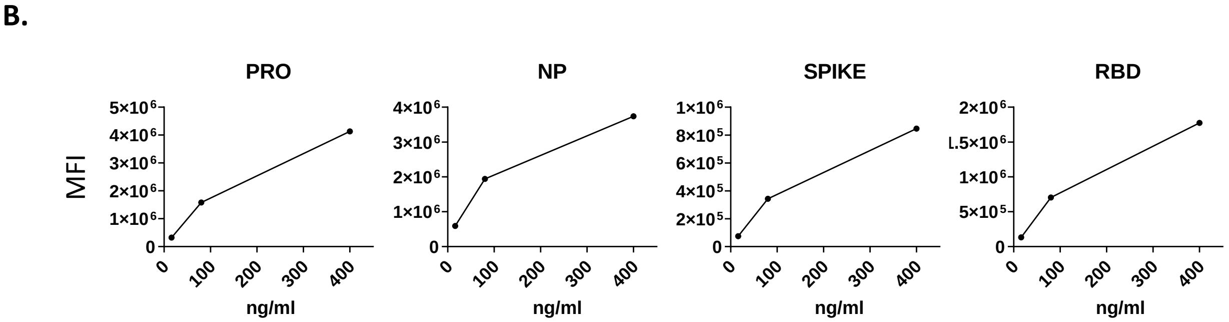


**Supplementary Figure 1. Titration of the anti-His antibody binding to individual viral antigen-coated beads. A. Estimation of the antibody-binding capacity of MPro beads.** Magnetic beads coated with SARS-CoV-2 MPro were incubated with the indicated amounts of anti-his-tag antibody, followed by anti-rabbit-PE and analysed by flow cytometry. The histogram shows the fluorescence intensity for each anti-His concentration. **B. Estimation of the antibody-binding capacity of SARS-CoV-2 antigen-coated beads**. The same was done in parallel with different magnetic beads, each one coated with one SARS-CoV-2 antigen: Spike, the Receptor Binding Domain (RBD) of the S, the nucleocapsid protein (NP) and 3CL main protease (Mpro). Three dilutions of anti-His antibody were used: 16 ng/ml, 80 ng/ml and 400 ng/ml. The plots represent the MFI values obtained for each condition tested.

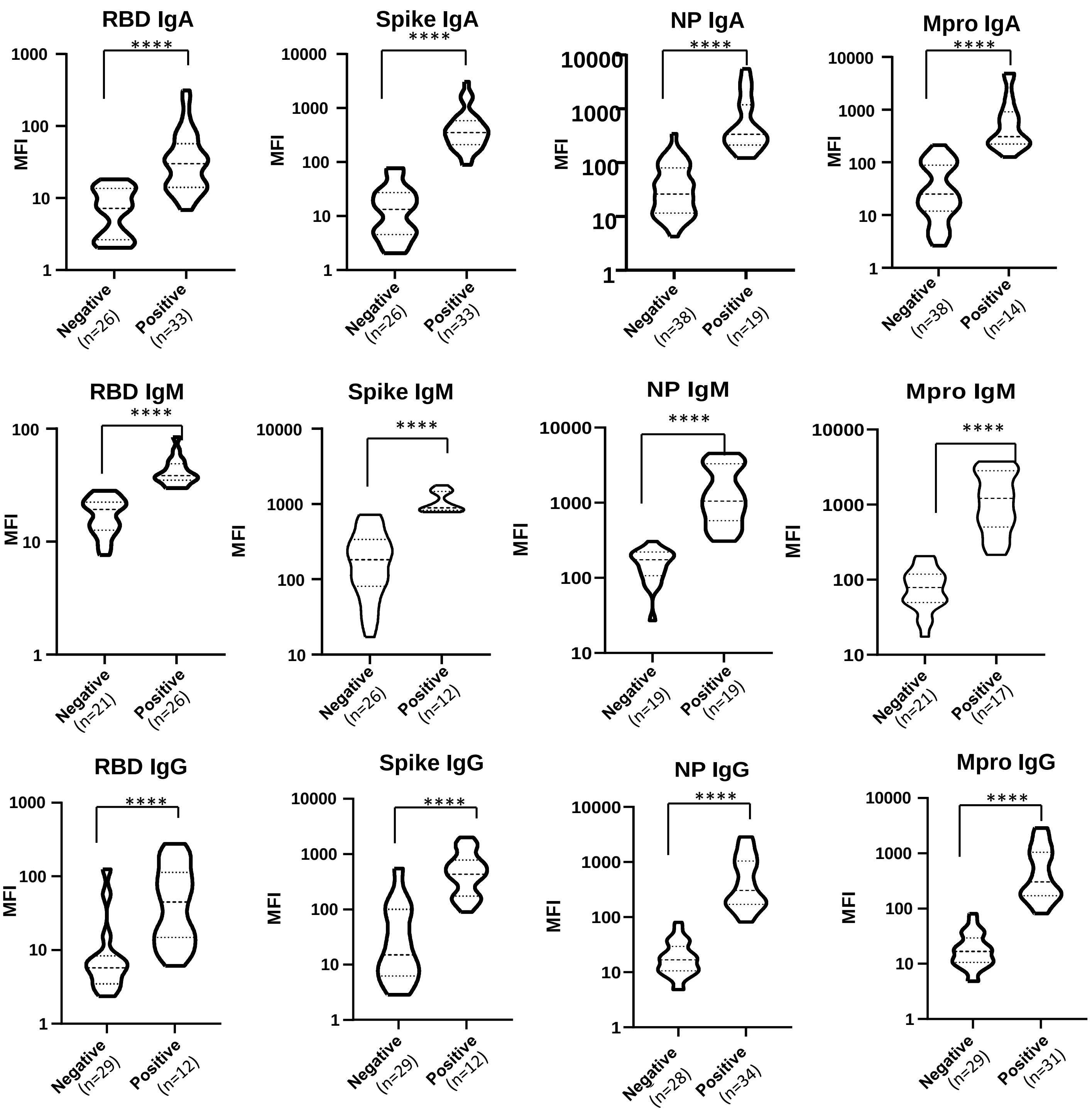


**Multiantigen assay for SARS-CoV-2 antibody detection.** Four different SARS-CoV-2 His-tagged antigens (Mpro, NP, S and RBD) were covalently coupled to magnetic beads labeled with dyes showing different fluorescence intensity in the APC and PerCP channels. Equal amounts of the different bead populations were mixed in the same tube and incubated with dilutions of plasma from patients or healthy donors, as indicated. Antibodies bound to the antigen were developed with fluorophore-conjugated anti-human Ig (FITC, PE and PE-Cyanine7) and samples were analysed by flow cytometry. Statistic comparison was carried out using a Mann-Whitney test.

**Supplementary Figure 3**

### A. IgG

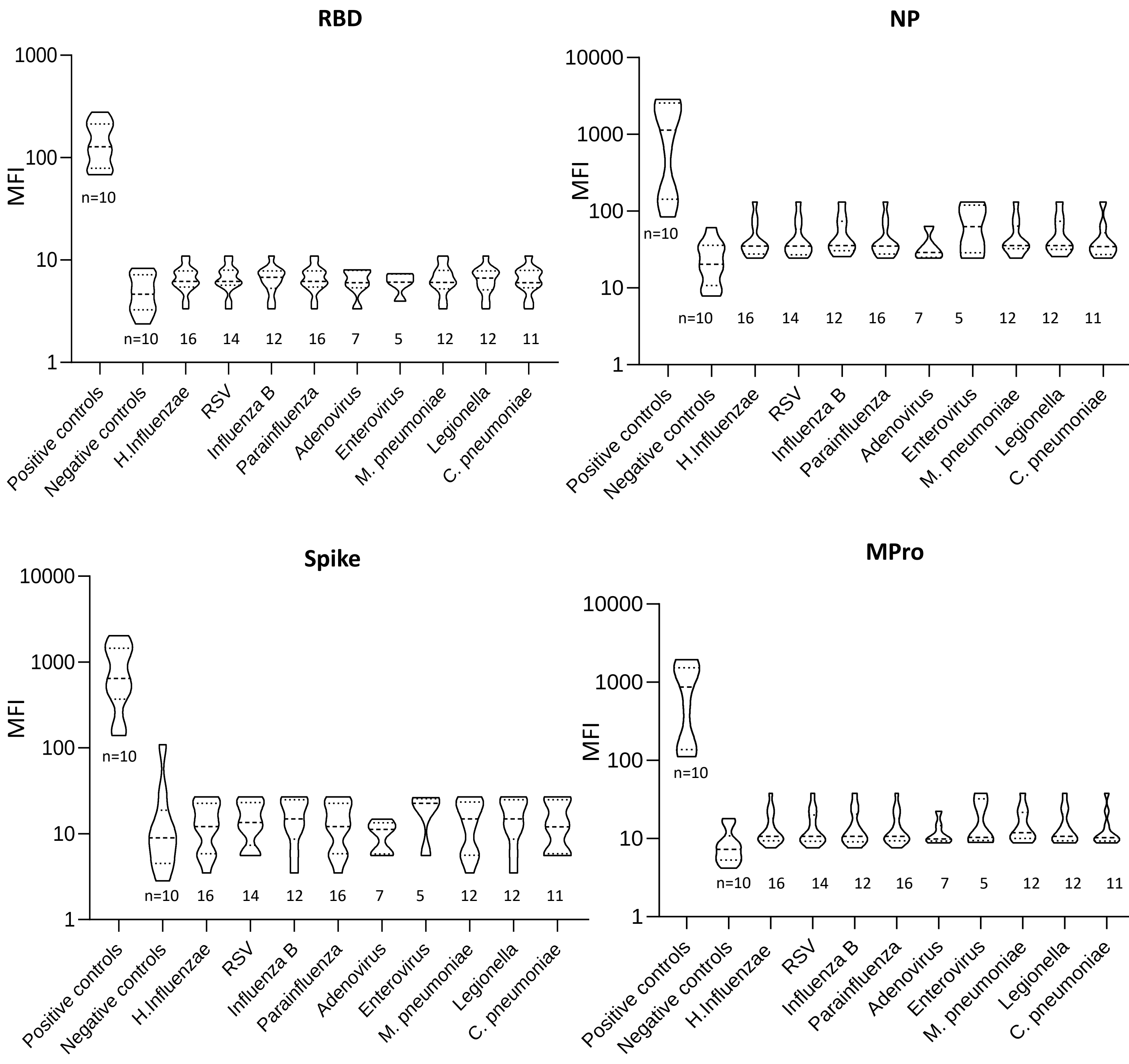

### B. IgA

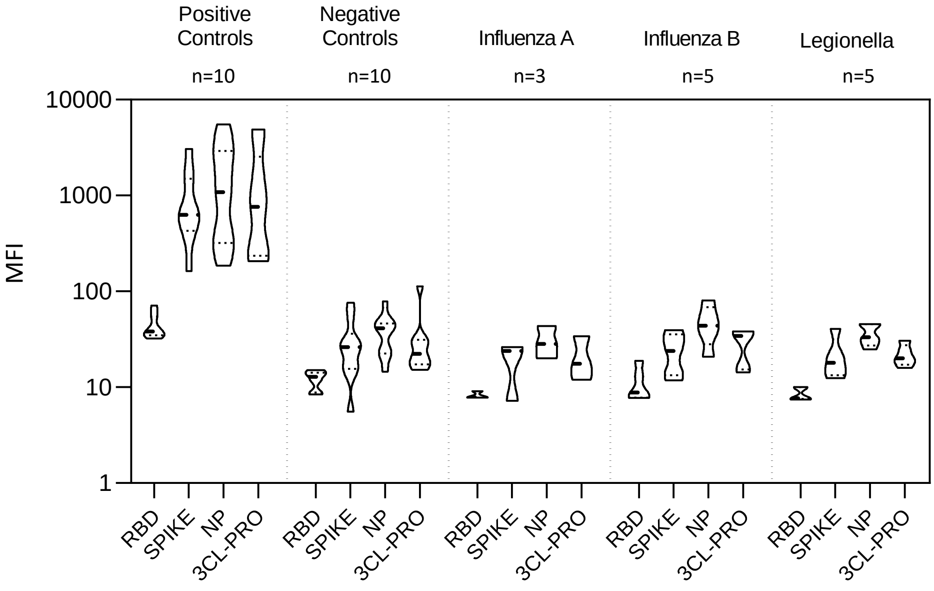


### C. IgM

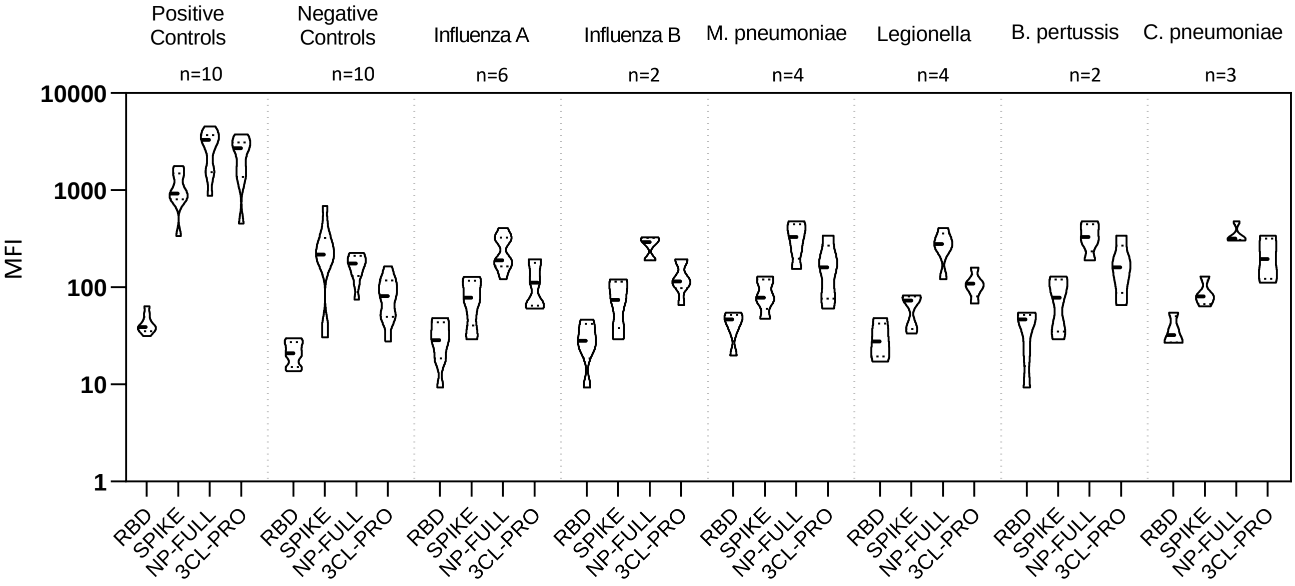


**Multiantigen assay for SARS-CoV-2 shows no cross-reacitivity with other respiratory diseases.** Four different SARS-CoV-2 His-tagged antigens (Mpro, NP, S and RBD) were covalently coupled to magnetic beads labeled with dyes showing different fluorescence intensity in the APC and PerCP channels. Equal amounts of the different bead populations were mixed in the same tube and incubated with dilutions of plasma from patients or healthy donors, as indicated. Antibodies bound to the antigen were developed with fluorophore-conjugated anti-human Ig (FITC, PE and PE-Cyanine7) and samples were analysed by flow cytometry. **A. IgG. B. IgA. C. IgM**

**Supplementary Table 1. Patient demographic and clinical data for severity study**

| | | | N=29 | % |
|---|---|---|---|---|
| Gender | Male | | 25 | 86 |
| | Female | | 4 | 14 |
| Age | 40-60 | | 19 | 66 |
| | 61-70 | | 5 | 17 |
| | > 71 | | 5 | 17 |
| Time from symptoms onset to sample collection | < 15 days | | 3 | 10 |
| | 15-30 days | | 8 | 28 |
| | 31-45 days | | 4 | 14 |
| | > 45 | | 9 | 48 |
| Hospitalization | Yes | ICU | 15 | 55 |
| | No | | 14 | 45 |
| Fever | | | 16 | 55 |
| Rhinorrhea | | | 1 | 3 |
| Anosmia | | | 4 | 14 |
| Ageusia | | | 3 | 10 |
| Odynophagia | | | 2 | 7 |
| Dry cough | | | 11 | 38 |
| Productive cough | | | 3 | 10 |
| Dyspnea | | | 11 | 38 |
| Pneumonia | | | 13 | 45 |
| Fatigue, myalgia, anorexia | | | 14 | 48 |
| Pulmonary infiltrate | | | 13 | 45 |
| Gastrointestinal symptoms | | | 0 | 0 |
| Cutaneous lesions | | | 0 | 0 |
| Multiorgan dysfunction | | | 5 | 17 |
| Thrombotic events | | | 2 | 7 |
| Comorbidities (HTN, DM, COPD, obesity, cancer) | | | 7 | 24 |
| Required mechanical ventilation | | | 13 | 45 |
| Death | | | 2 | 7 |

ICU (intensive care unit), HTN (hypertension), DM (diabetes mellitus), COPD (chronic obstructive pulmonary disease).

**Supplementary Table 2. Patient demographic for vaccination study**

| | | N=15 | % |
|---|---|---|---|
| Gender | Male | 5 | 33.3 |
| | Female | 10 | 66.6 |
| Age (25-56) | 25-35 | 4 | 26.6 |
| | 36-45 | 4 | 26.6 |
| | > 46 | 7 | 46.6 |
| Days from first dose (2-35) | 2 days | 1 | 6.6 |
| | 10-20 days | 5 | 33.3 |
| | 21-30 days | 6 | 40.0 |
| | > 31 | 3 | 20.0 |
| Patients having received 2nd dose | Y | 9 | 60.0 |
| | N | 6 | 40.0 |

**Supplementary Table 3. Prediction accuracy of healthy vs COVID samples***

| Samples | | ELISA | | | | FACS Multiantigenic | FACS RBD-Spike-Pro | | | |
|---|---|---|---|---|---|---|---|---|---|---|
| **Donor** | **Status** | **RBD** | **Spike** | **NP** | **Pro** | **1:100-1:800** | **1:100** | **1:200** | **1:600** | **1:1800** |
| **8** | Covid-19 | 100 | 100 | 100 | 100 | 100 | 100 | 100 | 100 | 100 |
| **11** | Covid-19 | 100 | 100 | 100 | 100 | 100 | 100 | 100 | 100 | 100 |
| **14** | Covid-19 | 100 | 100 | 100 | 100 | 100 | 100 | 100 | 100 | 100 |
| **15** | Covid-19 | 100 | 100 | 100 | 100 | 100 | 100 | 100 | 100 | 100 |
| **16** | Covid-19 | 100 | 100 | 100 | 100 | 100 | 100 | 100 | 100 | 100 |
| **18** | Covid-19 | 100 | 100 | 100 | 100 | 100 | 100 | 100 | 100 | 100 |
| **21** | Covid-19 | 100 | 100 | 100 | 100 | 100 | 100 | 100 | 100 | 100 |
| **97** | Covid-19 | 100 | 100 | 100 | 100 | 100 | 100 | 100 | 100 | 100 |
| **102** | Covid-19 | 100 | 100 | 100 | 100 | 100 | 100 | 100 | 100 | 100 |
| **103** | Covid-19 | 100 | 100 | 100 | 100 | 100 | 100 | 100 | 100 | 100 |
| **106** | Covid-19 | 100 | 100 | 100 | 100 | 100 | 100 | 100 | 100 | 100 |
| **107** | Covid-19 | 100 | 100 | 100 | 100 | 100 | 100 | 100 | 100 | 100 |
| **109** | Covid-19 | 100 | 100 | 100 | 100 | 100 | 100 | 100 | 100 | 100 |
| **110** | Covid-19 | 100 | 100 | 100 | 100 | 100 | 100 | 100 | 100 | 100 |
| **128** | Covid-19 | 100 | 100 | 11,54 | 89,87 | 100 | 100 | 100 | 100 | 100 |
| **35** | Covid-19 | 100 | 100 | 100 | 100 | 100 | 100 | 100 | 100 | 100 |
| **57** | Covid-19 | 47,43 | 99,29 | 0 | 13,02 | 99,93 | 98,46 | 100 | 99,78 | 99,92 |
| **92** | Covid-19 | 100 | 100 | 100 | 100 | 100 | 100 | 100 | 100 | 100 |
| **64** | Covid-19 | 100 | 99,92 | 11,72 | 24,81 | 100 | 100 | 100 | 99,7 | 100 |
| **99** | Covid-19 | 100 | 100 | 100 | 100 | 100 | 100 | 100 | 100 | 100 |
| **100** | Covid-19 | 100 | 100 | 100 | 100 | 100 | 100 | 100 | 100 | 100 |
| **101** | Covid-19 | 100 | 100 | 100 | 100 | 100 | 100 | 100 | 100 | 100 |
| **129** | Covid-19 | 100 | 100 | 100 | 100 | 100 | 100 | 100 | 100 | 100 |
| **114** | Covid-19 | 99,37 | 98,68 | 100 | 100 | 98,24 | 97,53 | 99,57 | 68,26 | 99,49 |
| **89** | Covid-19 | 96,46 | 100 | 100 | 100 | 100 | 100 | 100 | 100 | 100 |
| **115** | Covid-19 | 100 | 100 | 100 | 100 | 100 | 100 | 100 | 100 | 100 |
| **121** | Covid-19 | 100 | 100 | 100 | 100 | 100 | 100 | 100 | 100 | 100 |
| **123** | Covid-19 | 100 | 100 | 100 | 100 | 100 | 100 | 100 | 100 | 100 |
| **130** | Covid-19 | 100 | 100 | 100 | 100 | 100 | 100 | 100 | 100 | 100 |
| **43** | Control | 100 | 100 | 100 | 100 | 100 | 100 | 100 | 100 | 100 |
| **36** | Control | 100 | 100 | 100 | 100 | 100 | 100 | 100 | 100 | 100 |
| **187** | Control | 100 | 100 | 100 | 100 | 100 | 100 | 100 | 100 | 100 |
| **94** | Control | 100 | 100 | 100 | 100 | 100 | 100 | 100 | 100 | 100 |
| **24** | Control | 100 | 100 | 100 | 100 | 100 | 100 | 100 | 100 | 100 |
| **32** | Control | 100 | 100 | 100 | 100 | 100 | 100 | 100 | 100 | 100 |
| **33** | Control | 100 | 100 | 100 | 100 | 100 | 100 | 100 | 100 | 100 |
| **75** | Control | 100 | 100 | 100 | 100 | 100 | 100 | 100 | 100 | 100 |
| **2** | Control | 100 | 100 | 100 | 100 | 100 | 100 | 100 | 100 | 100 |
| **8** | Control | 100 | 100 | 100 | 100 | 100 | 100 | 100 | 100 | 100 |
| **109** | Control | 100 | 100 | 100 | 100 | 100 | 100 | 100 | 100 | 100 |
| **125** | Control | 100 | 100 | 2,29 | 1,11 | 100 | 100 | 100 | 100 | 100 |
| **112** | Control | 100 | 100 | 100 | 100 | 100 | 100 | 100 | 100 | 100 |
| **80** | Control | 100 | 100 | 100 | 100 | 100 | 100 | 100 | 100 | 100 |
| **29** | Control | 100 | 100 | 100 | 100 | 100 | 100 | 100 | 100 | 100 |

*Samples were stratified and randomly spliced into training and test sets. The training samples were used to fit a random forest classifier which then predicted the healthy vs disease category of unseen test samples (1/7 of total samples). This was repeated n=10,000 times. For each patient, accuracy was calculated as the proportion of correct predictions divided by the number of predictions made
